# Plasma levels of soluble podoplanin are higher in acute promyelocytic leukemia compared to other forms of acute myeloid leukemia

**DOI:** 10.1101/2025.07.29.25332218

**Authors:** Carla Roberta Peachazepi Moraes, Camilla Maria de Alencar Saraiva, Ivanio Teixeira de Borba-Junior, Bruno Kosa Lino Duarte, Paula Melo de Campos, Sara Teresinha Olalla Saad, Erich Vinicius De Paula

## Abstract

**Background:** acute promyelocytic leukemia (APL) is a subtype of acute myeloid leukemia (AML) marked by a high incidence of coagulopathy. Podoplanin (PDPN), a glycoprotein involved in platelet activation through interaction with CLEC-2, has been recently identified on leukemic promyelocytes and suggested as a potential contributor to APL coagulopathy. Identification of novel biomarkers and therapeutic targets for APL coagulopathy can potentially improve the outcomes of this condition.

**Aim:** to explore whether levels of soluble PDPN (sPDPN) in plasma are different in APL, and to evaluate its association with laboratory and clinical outcomes in these patients.

**Methods:** samples were obtained from consecutive patients with APL at the time of diagnosis in an academic hospital. Biobank samples from 35 patients with non-APL AML matched for age and sex were used as comparators. Circulating PDPN levels were measured in plasma using a commercial ELISA. The study was approved by the IRB and all participants provided written informed consent.

**Results:** APL patients showed significantly higher plasma sPDPN concentrations compared non-APL AML. Using the median sPDPN value as a cutoff, a higher proportion of APL patients presented elevated levels. sPDPN levels correlated with CD40L among APL cases, but not among non-APL AML patients, suggesting a possible interaction in with thrombo-inflammatory activation pathways.

**Conclusion:** these findings represent a proof-of-concept that measuring sPDPN in plasma samples can contribute to the diagnosis of APL, while also providing novel data on the association of PDPN with the pathogenesis of APL coagulopathy.

## Introduction

Acute promyelocytic leukemia (APL) is a distinct subtype of acute myeloid leukemia (AML) characterized by the accumulation of promyelocytes in the bone marrow and a high risk of severe hemorrhagic complications. Despite significant therapeutic advancements with all-trans retinoic acid (ATRA) and arsenic trioxide (ATO)(1–3), early mortality remains a major concern, potentially reaching 5-20%, depending on clinical and geographical factors. The primary cause of early death is coagulopathy, which often leads to fatal intracranial hemorrhages (4). Therefore, early diagnosis and prompt initiation of treatment are crucial for reducing mortality and improving patient outcomes. In addition, the identification of novel biomarkers and therapeutic targets of APL coagulopathy could bring benefits to these patients.

Podoplanin (PDPN) is a small transmembrane glycoprotein expressed in various cell types, including lymphatic endothelial cells, kidney podocytes, and certain tumor cells (5). Functionally, PDPN interacts with the C-type lectin-like receptor 2 (CLEC-2) on platelets, triggering their activation and contributing to thrombotic processes (6). This mechanism is particularly relevant in cancer-associated thrombosis and inflammatory diseases, where PDPN overexpression has been linked to prothrombotic states (5–7).

In the context of APL, PDPN expression was first identified in leukemic promyelocytes in 2018, suggesting its potential role as a distinguishing marker for this leukemia subtype (8). More recently, we validated its expression in a prospective cohort using flow cytometry, reinforcing its diagnostic and pathophysiological relevance in APL (9). However, the extent to which circulating PDPN correlates with disease characteristics and coagulation abnormalities remains unexplored.

Soluble forms of membrane proteins can serve as biomarkers, reflecting their biological activity and disease status. For instance, soluble fms-like tyrosine kinase-1 (sFLT1) is well established as a biomarker in preeclampsia and soluble urokinase-type plasminogen activator receptor (suPAR) as a biomarker in different types of cancers (10– 13). Similarly, soluble PDPN (sPDPN) has been identified as a biomarker in malignancies and inflammatory diseases such as cancer and COVID-19, highlighting its potential and prognostic significance (14–18).

Based on this evidence, we investigated whether quantification of sPDPN in plasma could be associated with the diagnosis of APL, as well as its relationship with biomarkers involved in APL coagulopathy.

## Methods

### Study population

This study analyzed patients diagnosed with acute leukemia between 2014 and 2022 at an academic tertiary hospital. Inclusion criteria were: age ≥18 years and a confirmed diagnosis of APL or AML. Exclusion criteria included: presence of severe infection with hemodynamic instability requiring vasoactive agents at the time of sample collection, or a diagnosis of other forms of acute leukemia. Patients with non-APL AML were used as a comparator group. These patients were selected from the population of consecutive new AML cases during the same study period, matching for age, sex and year of enrollment, before any of the laboratory analysis. The diagnostic workup comprised morphological evaluation, immunophenotyping, cytogenetic analysis, and molecular testing. APL cases were confirmed through molecular detection of the PML-RARA fusion gene. Patients with a strong clinical and morphological suspicion of APL, based on peripheral blood smear findings, were promptly initiated on ATRA therapy in accordance with institutional protocol. The study was approved by the institutional ethics committee (CAAE: 39948520.8.1001.5404) and performed in accordance with the Declaration of Helsinki.

### Sample collection and processing

Whole blood samples were collected in EDTA tubes. Plasma was obtained by centrifugation at 2,500g for 15 minutes at 22°C within 2 hours of collection. All samples were aliquoted and stored at -80°C until analysis.

### Clinical and laboratory data

Clinical and laboratory data were retrospectively retrieved from the hospital’s electronic medical records. Laboratory variables included initial bone marrow blast percentage, hemoglobin concentration, white blood cell (WBC) count, platelet count, prothrombin time (TP), activated partial thromboplastin time (aPTT), and fibrinogen concentration, based on the first set of results available upon hospital admission. Clinical outcomes evaluated comprised early mortality (defined as death within 30 days of diagnosis) (19) and central nervous system (CNS) and retinal bleeding.

### Measurement of soluble podoplanin levels

Plasma levels of podoplanin were measured in duplicate in EDTA plasma using a commercial enzyme-linked immunosorbent assay kits (ELISA) (Sigma Aldrich, Cat. RAB 1632-1KT - Lot: 0325/2117), according to the manufacturer’s instructions.

### Measurement of P-selectin and CD40L

Plasma concentrations of P-selectin and CD40L were determined using a custom-designed multiplex assay panel (Invitrogen – Thermo Fisher Scientific – Cat. PPX-07-MX323GU – Lot: 283814-000), following the manufacturer’s protocol. All measurements were performed in duplicate, and analyte concentrations were calculated based on standard curves generated using known concentrations of recombinant proteins.

### Statistical analysis

Quantitative variables were described using both median with interquartile range and mean with standard deviation (SD). Patients with APL were matched 1:1 to non-APL AML patients, as detailed above. Matching was done prior to any laboratory analysis. Comparative analyses between groups were performed using parametric and nonparametric statistical methods. For normally distributed data, the Student’s t-test was applied, while the Mann–Whitney U test was used for data not meeting parametric assumptions. For the analysis of the association between PDPN expression with clinical outcomes, APL patients were arbitrarily categorized as PDPN positive or negative, based on the 50th percentile cutoff. Correlations were evaluated by Spearman’s test. A P value < 0.05 was considered statistically significant. Statistical analyses were conducted using IBM SPSS Statistics version 26 and GraphPad Prism version 8.0.

## Results

In total, 35 patients with APL were included in the study, as well as 35 patients with non-APL AML used as a comparator group, matched by age, sex and year of diagnosis. The clinical and laboratory characteristics of these groups are presented in table 1. As expected, APL patients presented more frequently alterations in hemostasis biomarkers, and a lower early mortality.

**Table 1.** Laboratory and clinical characteristics of study participants.

|  | **APL** | **Non-APL AML** | **p<sup>‡</sup>** |
| --- | --- | --- | --- |
|  | **(n=35)** | **(n=35)** |  |
| Age* | 37.3 (21-61) | 41.4 (18-59) | 0.31 |
| Male:Female (ratio) | 17:18 | 17:18 | 0.99 |
| Risk group (low/int/high) | 3/18/14 | 11/11/13 | 0.04 |
| Peripheral blast count, 10<sup>9</sup>/L* | 2.23 (0.01-129.6) | 7.75 (0.1-130.8) | 0.25 |
| Bone marrow blast count, %* | 88.0 (46.0-98.2) | 80.5 (26.5-95.6) | 0.002 |
| Leukocytes, 10<sup>3</sup>/μL* | 3.1 (0.32-137.9) | 14.4 (0.91-150.3) | 0.02 |
| Hemoglobin, g/dL** | 8.4 ± 2.39 | 8.1 ± 2.10 | 0.64 |
| Platelets, 10<sup>9</sup>/L* | 22.0 (4.0-89.0) | 27.0 (5.0-230.0) | 0.02 |
| PT, INR** | 1.29 ± 0.19 | 1.19 ± 0.13 | 0.01 |
| aPTT, INR* | 0.90 (0.75-1.24) | 0.89 (0.67-1.38) | 0.48 |
| Fibrinogen, mg/dL* | 121.7 (26.0-378.5) | 384.5 (58.9-705.8) | <0.0001 |
| ***Clinical outcomes*** |  |  |  |
| CNS/retinal bleeding, n (%) | 4 (11.4) | 2 (5.7) | 0.39 |
| 30-day survival (no/yes) | 5/30 | 12/23 | 0.05 |

\*Median (interquartile range); \*\*mean ± SD; ‡Mann-Whitney test, t-test or chi-square test; APL: acute promyelocytic leukemia; AML: acute myeloid leukemia; CNS: central nervous system; prothrombin time (PT); activated partial thromboplastin time (aPTT).

Plasma concentrations of sPDPN ranged from 0.3 to 113.6 ng/mL. Patients with APL exhibited significantly higher levels of sPDPN (median= 16.4 ng/mL) compared to patients with other AML subtypes (median= 2.2 ng/mL) (p = 0.003) (figure 1).

**Figure 1.**
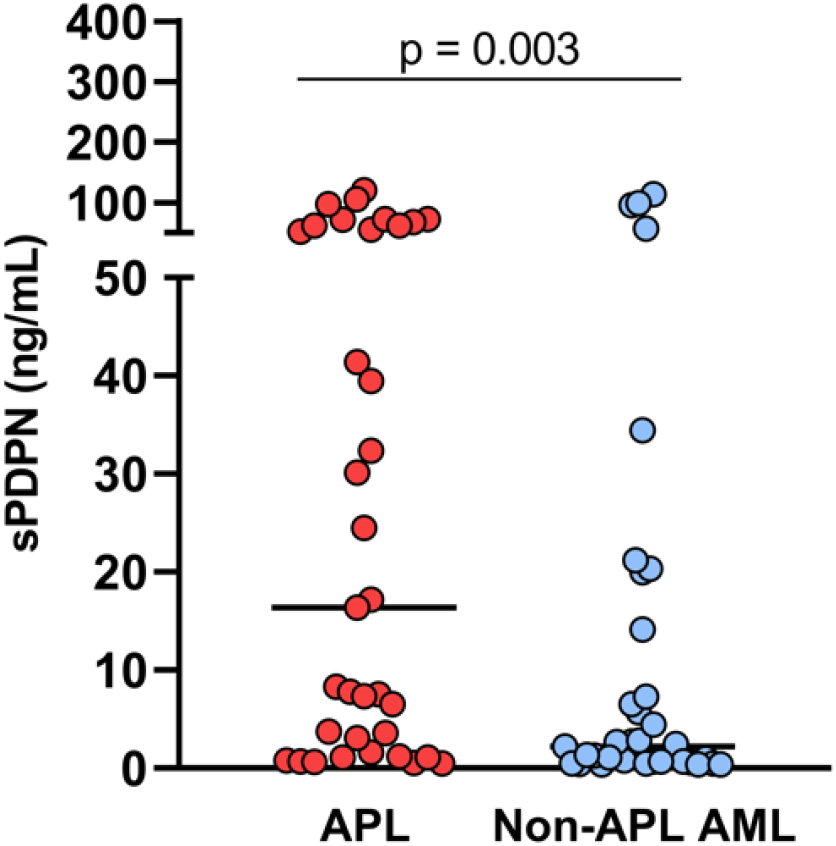
(A) Plasma levels of PDPN among patients with APL and non-APL AML, P from Mann-Whitney test.

To investigate the potential relationship between sPDPN and platelet activation, we measured plasma levels of P-selectin and CD40L, two established markers of platelet degranulation. Both P-selectin (7,194 ng/mL in APL vs. 11,220 ng/mL in non-APL AML, p = 0.003) and CD40L levels (100.4 ng/mL in APL vs. 196.6 ng/mL in non-APL AML, p = 0.002) were significantly lower in APL patients compared to those with other AML subtypes (Figure 2A–B).Interestingly, a significant positive correlation was observed between PDPN and CD40L levels among APL patients (R_s_ = 0.60; p = 0.0004), but not among other AML subtypes (figure 2C-D). PDPN levels were not associated with predefined clinical outcomes (table 2).

**Table 2.** Association of PDPN expression with clinical outcomes in APL.

| *Clinical outcome* | *PDPN expression category\** |  | *p\*\** |
| --- | --- | --- | --- |
|  | PDPN<sup>+</sup> | PDPN<sup>-</sup> |  |
| Early mortality (30-day) |  |  | 0.46 |
| Yes | 4 | 1 |  |
| No | 19 | 11 |  |
| CNS/ retinal bleeding |  |  | 0.67 |
| Yes | 3 | 1 |  |
| No | 20 | 11 |  |

\*For the purpose of association analyses, APL patients were arbitrarily categorized as PDPN positive or negative, based on the 50th percentile cutoff. \*\*Statistical comparison was performed using the chi-square test.

**Figure 2.**
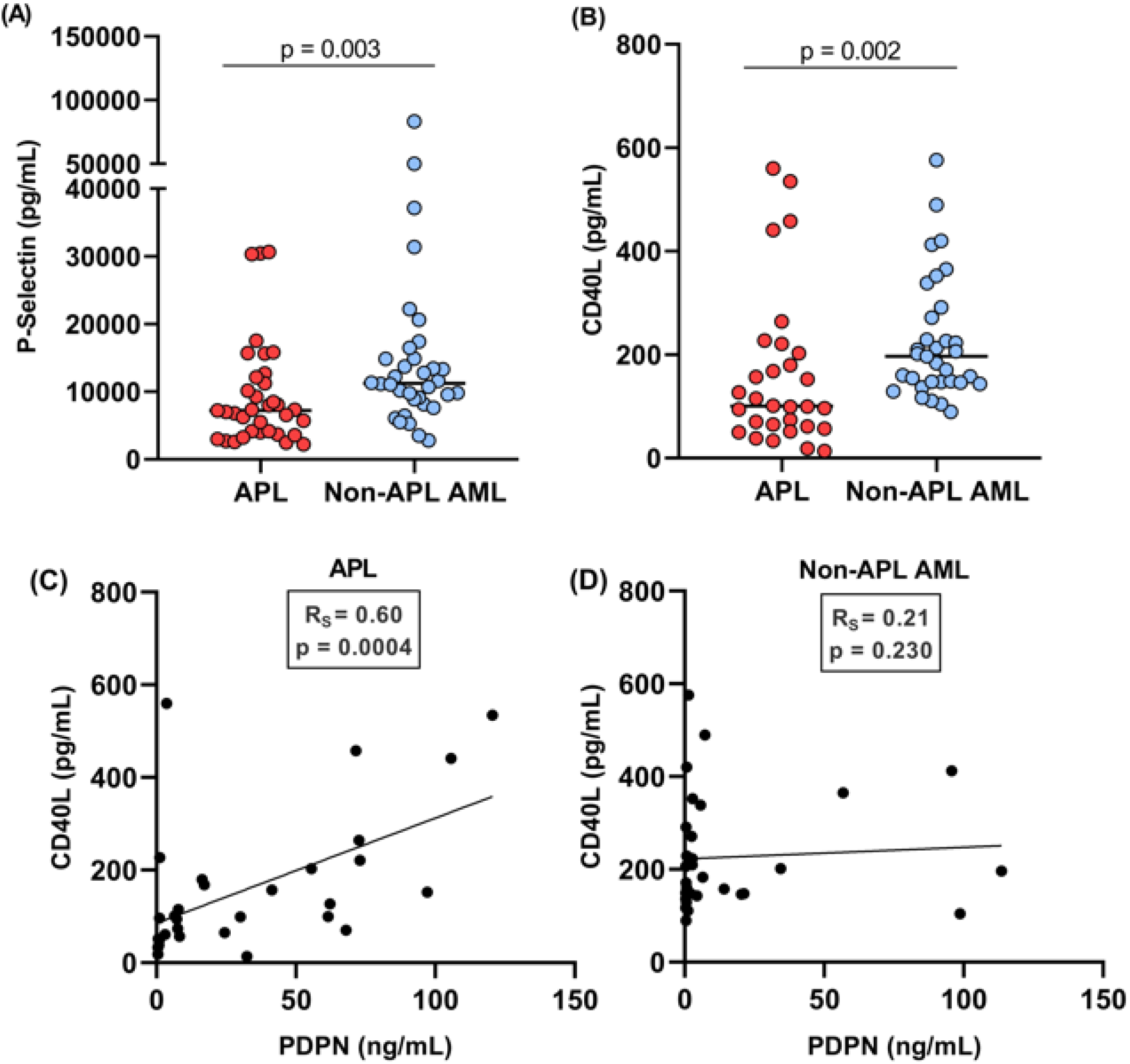
(A) Plasma levels of P-selectin were significantly lower in APL patients compared to non-APL AML patients (Mann–Whitney test; p = 0.003). (B) Plasma levels of CD40L were significantly lower in APL patients compared to non-APL AML patients (Mann–Whitney test; p = 0.002). (C) A significant positive correlation between PDPN and CD40L was observed in APL patients (Spearman’s correlation; p = 0.0004). (D) No significant correlation between PDPN and CD40L was found in non-APL AML patients.

## Discussion

APL coagulopathy still represents a major challenge to the early management of APL, and the availability of biomarkers capable to differentiate APL from other forms of AML, and to provide information about APL coagulopathy could potentially facilitate the management of these patients (20). The main contribution of our study was the demonstration of sPDPN as a potential biomarker whose expression is markedly higher in APL compared to other forms of AML, and that also correlates with an important thrombo-inflammatory mediator, thus suggesting that PDPN expression may be a unique biological feature that contributes to the well-known predisposition of these patients to hemorrhagic and thrombotic complications.

The first demonstration that PDPN expression is a hallmark of APL was published in 2018 and included data obtained by flow cytometry and RNA sequencing (8). That study also presented evidence linking PDPN expression to platelet activation *in vitro*. While the diagnosis of APL using flow cytometry and molecular techniques is well established, reliance on methods that are not usually available outside specialized cancer centers may delay diagnosis and jeopardize patient outcomes (20). Immune-based method such as ELISA are widely available and have the advantage of being potentially performed with diagnostic kits in almost every setting. Based on the demonstration that circulating levels of PDPN was capable to identify discrete subgroups of patients in other conditions (14–18), and that soluble levels of membrane proteins can serve as useful disease biomarkers (10–13), we hypothesized that sPDPN levels might contribute to the differential diagnosis of APL with other forms of AML. In fact, we were able to demonstrate a significant difference in sPDPN levels between these two patient subgroups, although a subgroup of patients with non-APL AML also presents higher sPDPN values.

In addition, we found a positive correlation between sPDPN and CD40L levels in APL patients, suggesting a biological interaction between these two markers. CD40L is a transmembrane protein expressed on activated platelets that has been implicated in platelet-leukocyte crosstalk and inflammatory signaling (21). CD40L is also expressed by other hematopoietic cells such as lymphocytes, neutrophils, dendritic cells, monocytes and macrophages, in response to cytokines (22), which are key elements of thrombo-inflammatory pathways (23). So, the correlation described in our study could represent either evidence of the participation of PDPN in platelet activation, in alignment with Lavallé *et al* (8), who demonstrated that PDPN-expressing APL blasts can activate platelets, or a yet unknown association between PDPN and CD40L in another hematopoietic compartment. In our dataset, sPDPN levels were not associated with clinical outcomes, although our study was not powered for this type of analysis.

Our results also demonstrate that a significant part of AML patients also present detectable sPDPN levels, while some APL patients present lower sPDPN levels. This is in contrast with data from flow cytometry and RT-PCR from our lab that show less overlap between APL and other forms of AML (data not shown). Additional studies are warranted to explore and explain this lower accuracy of sPDPN ELISA for the segregation of APL from other forms of AML compared to flow cytometry.

Our study has limitations that should be acknowledged. Although APL is a relatively rare condition and our sample size was sufficient to demonstrate relevant biological differences in sPDPN levels, our study was not powered to analyze clinical outcomes, which were rare in our population. Another limitation lies in the absence of additional hemostatic biomarkers, which might have enhanced the interpretation of coagulation-related mechanisms. However, these markers are not routinely assessed in the hospital’s standard clinical workflow, and citrate plasma samples were no longer available. Future studies involving larger cohorts and broader biomarker panels will be important to further investigate the potential diagnostic and clinical relevance of sPDPN in APL.

In conclusion, our results represent a proof of concept that the immunodetection (in this case, using ELISA) of circulating levels of sPDPN can serve as a complementary tool in the differential diagnosis of APL. Additionally, our data also adds support to the concept that PDPN expression is a distinct hallmark of APL and contributes to the pathogenesis of the unique thrombo-inflammatory complications seen in these patients.

## Data Availability Statement

All reported data are available for sharing upon a reasonable request to the corresponding author.

## Ethics Statement

The study was performed in accordance with the Declaration of Helsinki and approved by the local IRB (protocol CAAE: 39948520.8.1001.5404).

## Author contributions statement

CRPM: performed all assays; revised medical records; contributed to data analysis and drafted the manuscript; revised and approved the manuscript. CMAS: revised medical records; contributed to data analysis; revised and approved the manuscript. ITBJ: contributed to data analysis; revised and approved the manuscript. BKLD: contribute to study design; revised and approved the manuscript. PMC: provided laboratory support and infra-structure; revised and approved the manuscript. STOS: provided laboratory support and infra-structure; revised and approved the manuscript. EVDP: designed the study, oversaw and provided resources and infra-structure for ELISA analysis, contributed to data analysis; drafted the manuscript; revised and approved the manuscript.

## Acknowledgements

The author(s) disclosed receipt of the following financial support for the research, authorship, and/or publication of this article: This study was funded by the Sao Paulo Research Foundation (FAPESP), grants 2020/05985-9, 2022/13216-0 and 2023/03765-0. FAEPEX-Unicamp grant 2404/2020 and 2633/23; Coordenacao de Aperfeicoamento de Pessoal de Nivel Superior – Brasil (CAPES), finance code 001.

